# Association Between Ultrasonography Fetal Anomalies and Autism Spectrum Disorder

**DOI:** 10.1101/2021.07.20.21260824

**Authors:** Ohad Regev, Amnon Hadar, Gal Meiri, Hagit Flusser, Analya Michaelovski, Ilan Dinstein, Reli Hershkovitz, Idan Menashe

## Abstract

**Background:** Prenatal ultrasound is frequently used to monitor fetal growth and identify fetal anomalies that may suggest genetic or developmental abnormalities which may develop into congenital anomalies and diseases. Autism spectrum disorder (ASD) is a complex neurodevelopmental disorder, associated with a wide range of congenital anomalies. Nevertheless, very little has been done to investigate organ development using prenatal ultrasound as a means to identify fetuses with ASD susceptibility.

**Methods:** A retrospective matched case-sibling-control study. ASD cases were matched to two control groups: *typically developing sibling* (TDS) closest in age to ASD child; and *typically developing population* (TDP), matched for age, sex, and ethnicity. The study comprised 659 children: 229 ASD, 201 TDS, and 229 TDP; 471 (71.5%) males.

**Results:** Ultrasonography fetal anomalies (UFAs) were found in 29.3% of ASD cases vs. only 15.9% and 9.6% in the TDS and TDP groups (aOR=2.23, 95%CI=1.32-3.78, and OR=3.50, 95%CI=2.07-5.91, respectively). Also, multiple co-occurring UFAs were significantly more prevalent among ASD cases. UFAs in the urinary system, heart, and head&brain were the most significantly associated with ASD diagnosis (aOR_Urinary_ =2.08, 95%CI=0.96-4.50 and aOR_Urinary_=2.90, 95%CI=1.41-5.95; aOR_Heart_=3.72, 95%CI=1.50-9.24 and aORHeart=8.67, 95%CI=2.62-28.63; and aOR_Head&Brain_=1.96, 95%CI=0.72-5.30 and aOR_Head&Brain_=4.67, 95%CI=1.34-16.24; vs. TDS and TDP, respectively). ASD females had significantly more UFAs than ASD males (43.1% vs. 25.3%, *p*=0.013) as well as higher prevalence of multiple co-occurring UFAs (15.7% vs. 4.5%, *p*=0.011), while no sex differences were seen among TDS and TDP controls. ASD fetuses were characterized by a narrower head and a relatively wider ocular-distance vs. TDP fetuses (aOR_BPD_=0.81, 95%CI=0.70-0.94, and aOR_Ocular-Distance_=1.29, 95%CI=1.06-1.57). Finally, UFAs were associated with more severe ASD symptoms.

**Conclusions:** Our findings shed important light on the abnormal multiorgan embryonic development of ASD and suggest fetal ultrasonography biomarkers for ASD.

## Introduction

Autism spectrum disorder (ASD) is a multifactorial, life-long neurodevelopmental disorder characterized by impaired social communication and restrictive-repetitive behaviors.^1,2^ However, many people with ASD manifest additional comorbidities and congenital anatomical abnormalities that further complicate their clinical picture.^3–16^ Nonetheless, today, the diagnosis of ASD is based on behavioral symptoms,^2^ which are typically manifested in the second year of life.^17^ A growing body of evidence suggests that the initial signs of ASD emerge during early childhood^18–20^ and possibly even before birth.^21–26^ Indeed, recent postnatal studies have found indications of the prenatal onset of abnormal neurodevelopment in children with ASD,^22^ and some prenatal studies have provided preliminary indications for abnormal brain development,^21,23,26^ and higher rates of structural anomalies in the renal system of both ASD fetuses and children with specific genetic syndromes associated with ASD.^27–32^ Taken together, these findings suggest that ASD may be associated with abnormal embryonic organogenesis of different body parts, which consequently leads to postnatal malformations in some children with ASD.^12,22,33,34^ Accordingly, there is emerging interest in examining the prenatal organ development of fetuses later developing into children diagnosed with ASD.^21,23,26^

Prenatal ultrasound, a commonly used pregnancy monitoring tool, allows physicians to survey fetal growth and organ development and may hence reveal anomalies suggesting genetic and developmental problems that require further testing and follow-up. In prenatal monitoring protocols, one of the primary ultrasound screenings is the fetal anatomy survey, which is considered standard of care for examining fetal organ development and detecting fetal organ anomalies. The survey involves the screening of the different organ systems and the measurement of a number of markers.^35,36^ The abnormalities that can be detected by the survey include structural anomalies and “soft markers” that may indicate genetic abnormalities or other non-genetic embryonic insults such as intrauterine infections, but some may be considered, in isolation, as normal variants or transient. The discovery of either structural abnormalities or “soft markers” during the fetal anatomy survey will usually prompt a thorough examination of the fetal anatomy and consideration of further diagnostic testing for chromosome abnormalities.^35–41^

Despite the emerging literature suggesting the prenatal onset of abnormal organogenesis and neurodevelopment in children with ASD and the possible genetic and environmental background of ASD, together with evidence of higher rates of congenital anomalies in ASD, very little has been done to investigate prenatal organ development in children with ASD, as reflected in the prenatal fetal anatomy survey. Specifically, all studies conducted to date only used basic biometric measures taken during the 2^nd^ and 3^rd^ trimesters, which do not allow thorough examination of fetal organ development.^21,23,26^ For this reason, we conducted the first study of ultrasound data from the fetal anatomy survey of fetuses developing into children later diagnosed with ASD in comparison with the ultrasound data for their unaffected siblings and for typically developing children from the general population.

## Materials and Methods

### Study Population

All the participants in this study were born between 2004 and 2018 to mothers living in southern Israel – the Negev – which has ∼700,000 inhabitants belonging to two main ethnic groups, Jews and Bedouins, that differ in their environmental exposures and genetic backgrounds. We included only fetuses from singleton pregnancies whose mothers were members of Clalit Health Services (CHS), Israel’s largest health maintenance organization (HMO), serving ∼75% of the Negev population. Members of CHS in this region receive most of their hospital-related health services (including ASD diagnosis) at the region’s only tertiary hospital, the Soroka University Medical Center, and its associated outpatient clinics.

### Study design

This retrospective case-sibling-control study comprised children diagnosed with ASD (cases); their own typically developing, closest-in-age siblings (TDS); and typically developing children from the [general] population (TDP), who were matched to cases by year of birth, sex (male/female) and ethnicity (Jewish/Bedouin). The case group was drawn from all children diagnosed with ASD in the Negev area, who are registered in the database of the National Autism Research Center of Israel (NARCI).^42,43^ The diagnosis of ASD at the NARCI is a multidisciplinary process, which entails a comprehensive intake interview (socio-demographic and clinical factors), a behavioral evaluation with ADOS-2,^44^ and a full neurocognitive assessment as described previously.^42,43^ The final diagnosis of ASD is made by a pediatric psychiatrist or neurologist, according to DSM-5 criteria.^2^

Of the 704 singleton birth children with ASD in the NARCI database (database freeze, February 2020), there were 237 children (34%) for whom the relevant ultrasound scans were available in the database of the CHS prenatal ultrasound clinics. Among the 237 children, there were eight pairs of siblings with ASD (multiplex families). We randomly assigned one ASD sibling from each such multiplex family to the final study sample to reduce familial bias in our results. In addition, a sensitivity analysis using the second ASD sibling from these families was conducted. In total, the study cohort included 659 children: 229 with ASD, 201 TDS, and 229 TDP (**Figure 1**). An evaluation of socio-demographic and clinical differences between cases in the study cohort and the other children with ASD in the NARCI database showed a lower proportion of Jews (vs. Bedouin Israelis), a lower parental age, and a higher ADOS score for the ASD children in the study cohort (**Supplementary Table S1**). The lower proportion of Jews in the study cases can be explained due to the more frequent use of private insurance among Jewish parents to conduct a more comprehensive anatomy survey than that offered by the HMO.^45,46^ The ethnic differences in the study cases can also explain the differences in parental age and ADOS scores, since Bedouins tend to have children at an earlier age than Jews,^43^ and the diagnosis of ASD in the Bedouin population is usually made for those with more severe symptoms of the disorder.^43,47^

**Figure 1.**
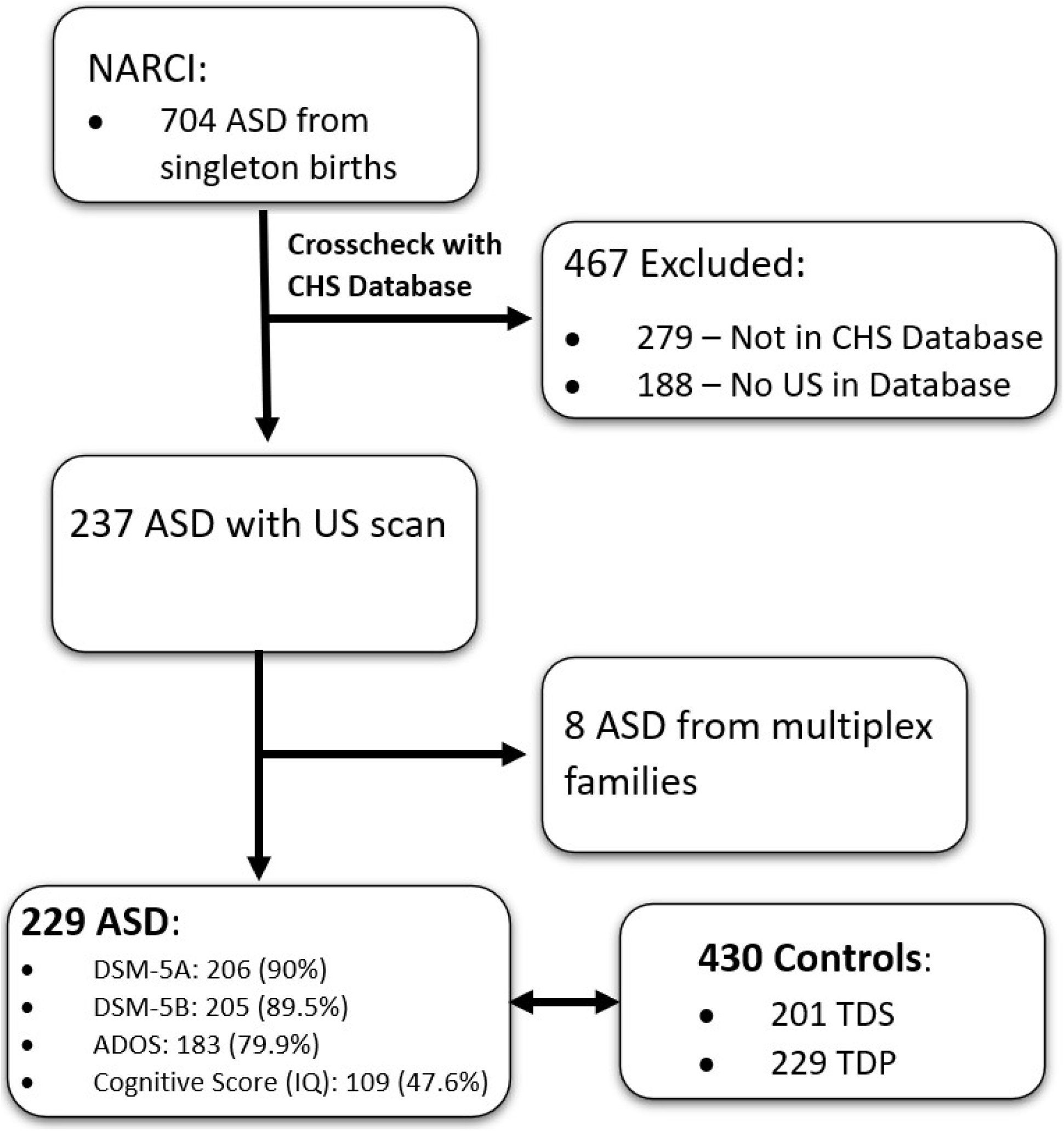
Flowchart of Children included in this Study.

### Fetal Ultrasound Data

Fetal ultrasound data from the fetal anatomy survey, which is conducted during gestational weeks 20-24 in Israel, were obtained from all the prenatal ultrasound clinics of CHS in southern Israel. In these clinics, fetal anatomy surveys are performed by experienced physicians, who record fetal anomalies and biometric measures according to standard clinical guidelines.^35,36^ The anatomy survey includes examination of different anatomical landmarks according to the various body systems, including the head, brain, thorax, abdomen, spine, limbs, and umbilical cord. Abnormalities in each examined organ are classified as either structural anomalies or “soft markers.”^35–38^ In addition, the following biometric measures are recorded: head circumference (HC), biparietal diameter (BPD), abdominal circumference (AC), femur length (FL), cisterna magna size, cerebellar diameter, lateral ventricle width, and ocular distance.^35,48^ The physician also assesses the fetal well-being according to a biophysical profile, which includes examination of the amniotic fluid index (AFI), breathing, movement, and tone, giving a score of 0–8.^49^ For the current study, the gestational age (GA) of each fetus was calculated from the last menstrual period (LMP) and confirmed by the crown-rump length (CRL) from the ultrasound scan in the first trimester. If the date of LMP was unknown, GA was calculated based on CRL.

### Statistical Analysis

We converted the basic biometric fetal measures (HC, BPD, AC, FL) to gestation-matched standardized Z-scores using the Hadlock approach, the most widely used standardization approach in this field.^50–52^ In addition, the proportions of the ocular distance and of the cerebellum width out of the BPD were calculated (e.g., ocular distance * 100/ BPD) in light of the strong relationships between these measures and head width. Differences in socio-demographic and clinical characteristics and in the proportion of anomalies between cases and each of the two matched control groups (TDS and TDP) were assessed using appropriate univariate statistics. Multivariable conditional regression or logistic regression models were used to assess the independent association of each ultrasound fetal measure/biomarker with ASD risk after adjusting for potential confounders. Finally, the association between clinical severity and fetal abnormalities was assessed using appropriate univariate statistics. P-values of analyses with multiple testing were adjusted using the Bonferroni correction. All analyses were conducted using SPSS Statistics V. 25 and R software. A two-sided test significance level of 0.05 was used throughout the entire study.

### Ethics Statement

The study was approved by the SUMC Ethics Committee per the Helsinki declaration SOR 295-18. Importantly, to protect patient confidentiality, all ultrasound data were ‘de-identified’ manner (i.e., without the mother’s ID or name, or any other identifiable information about the mother or the child).

## Results

### Socio-Demographic and Clinical Characteristics

Clinical and socio-demographic characteristics of the study sample are shown in **Table 1**. The anatomy survey was performed at the gestational age of 22.85 ±1.7 weeks, with no significant differences between the groups. Similarly, there were no significant differences between the groups in all other clinical characteristics, except for the inherent male bias in the ASD group compared to their unaffected siblings (77.7% vs. 56.7%, respectively; *p* <0.001).

**Table 1.** Clinical and Sociodemographic Characteristics for Children included in this Study.

| Variable |  | ASD<br>(n = 229) | TDS<br>(n = 201) | TDP<br>(n = 229) |
| --- | --- | --- | --- | --- |
| **Sociodemographic background** |  |  |  |  |
| Ethnicity (Jewish) | No. (%) | 163(71.2) | 141(70.1) | 163(71.2) |
|  | *P*<sup>a</sup> |  | 1 | 1 |
| Sex (male) | No. (%) | 178(77.7) | 114(56.7) | 178(77.7) |
|  | *P*<sup>a</sup> |  | **<0.001** | 1 |
| **Pregnancy Details** |  |  |  |  |
| Mother's age (years) | Mean±SD | 28.7±5.5 | 28.5±5.0 | 27.9±4.6 |
|  | *P*<sup>b</sup> |  | 1 | 0164 |
| Pregnancy number | Median (IQR) | 2(1-3) | 2(2-4) | 2(1-3) |
|  | *P*<sup>c</sup> |  | 0.242 | 0.234 |
| Previous abortions | Median (IQR) | 0(0-1) | 0(0-1) | 0(0-0) |
|  | *P*<sup>c</sup> |  | 1 | 0.070 |
| Gestational age at birth (weeks) | Mean±SD | 38.8±2.5 | 39.1±1.9 | 39.2±1.6 |
|  | *P*<sup>b</sup> |  | 0.496 | 0.188 |
| C section | No. (%) | 40(17.8) | 27(14.4) | 34(14.9) |
|  | *P*<sup>a</sup> |  | 0.696 | 0.818 |
| Birth weight (g) | Mean±SD | 3148 ±606 | 3163±593 | 3259±531 |
|  | *P*<sup>b</sup> |  | 1 | 0.128 |
| 1-min low APGAR score (<7) | No. (%) | 5(4.7) | 5(5.6) | 9(6.8) |
|  | *P*<sup>d</sup> |  | 1 | 0.966 |
| **US Details** |  |  |  |  |
| Gestation age assessed by last menstrual period | No. (%) | 157(76.2) | 130(75.6) | 166(82.2) |
|  | *P*<sup>a</sup> |  | 1 | 0.276 |
| Gestational age at US (weeks) | Mean±SD | 22.9±1.9 | 22.7±1.8 | 22.9±1.5 |
|  | *P*<sup>b</sup> |  | 0.412 | 1 |
| Placenta position, No. (%) | Fundus | 13(5.7) | 13(6.8) | 14(6.3) |
|  | Front wall | 124(54.6) | 102(53.7) | 109(48.7) |
|  | Back wall | 96(39.6) | 74(38.9) | 100(44.6) |
|  | Placenta previa | 0(0) | 1(0.5) | 1(0.4) |
|  | *P*<sup>a</sup> |  | 1 | 0.948 |
| Placental grading | Median (IQR) | 1(0-2) | 1(0-2) | 1(0-2) |
|  | *P*<sup>c</sup> |  | 1 | 0.578 |
| Breech Presentation at US | No. (%) | 74(32.9) | 58(29.6) | 59(26.1) |
|  | *P*<sup>a</sup> |  | 0.934 | 0.228 |
| Normal amniotic fluid | No. (%) | 222(97.4) | 195(98.5) | 219(97.8) |
|  | *P*<sup>d</sup> |  | 1 | 1 |
| Weight at US (g) | Mean±SD | 578±197 | 555±176 | 577±145 |
|  | *P*<sup>b</sup> |  | 0.406 | 1 |

Note: Boldface type indicates $p < 0.05$ . All p-values are Bonferroni corrected for multiple comparison (N=2). ASD = autism spectrum disorder; TDS = typically developing siblings; TDP = typically developing population; US = ultrasound.
<sup>a</sup> Chi-square; <sup>b</sup> Two-sided t-test; <sup>c</sup> Mann-Whitney U test; <sup>d</sup> Fisher's Exact Test

### Ultrasonography Fetal Anomalies (UFAs)

Case-control differences in UFAs are depicted in **Figure 2** and **Table 2**. Overall, UFAs were found in 67 (29.3%) of the ASD cases compared to 32 (15.9%) and 22 (9.6%) in the TDS and TDP groups (aOR=2.23, 95%CI=1.32-3.78, and aOR=3.50, 95%CI=2.07-5.91, respectively). In addition, more ASD cases had multiple anatomic anomalies than controls, with 7% of cases having multiple anatomic anomalies compared to only 2% of TDS and 0.9% of TDP controls (*p*=0.014 and *p*=0.001, respectively) (**Figure 2A**). Most UFAs in the ASD cases were seen in the urinary system and the heart (12.8% and 12.1%, respectively), followed by the head&brain (always taken together in this study; 5.7%), limbs (3.1%), blood vessels (2.7%), and gastrointestinal system (0.9%) (**Figure 2B**). Of note, cardiac UFAs were significantly associated with higher odds of an ASD diagnosis (aOR_Heart_=3.72, 95%CI=1.50-9.24, and aOR_Heart_=8.67, 95%CI=2.62-28.63 compared to TDS and TDP, respectively). UFAs in the urinary system were also associated, although to a lesser extent, with elevated odds of an ASD diagnosis (aOR_Urinary_ =2.08, 95%CI=0.96-4.50 and aOR_Urinary_=2.90, 95%CI=1.41–5.95 compared to TDS and TDP, respectively), with dilation of the renal pelvis (pyelectasis or hydronephrosis) being the most frequently detected anomaly. Finally, UFAs in the head&brain were significantly higher in the ASD cases compared to the TDP group but not to the TDS group (aOR_Head&Brain_=4.67, 95%CI=1.34-16.24, and aOR_Head&Brain_=1.96, 95%CI=0.72-5.30, respectively), with anomalies in CSF circulation seen among 4% of ASD cases compared to 1.3% of TDP (*p*=0.078). ASD cases also had higher rates of UFAs in most other organs examined in the study, although these differences did not reach statistical significance (**Table 2** and **Figure 2B**).

**Figure 2:**
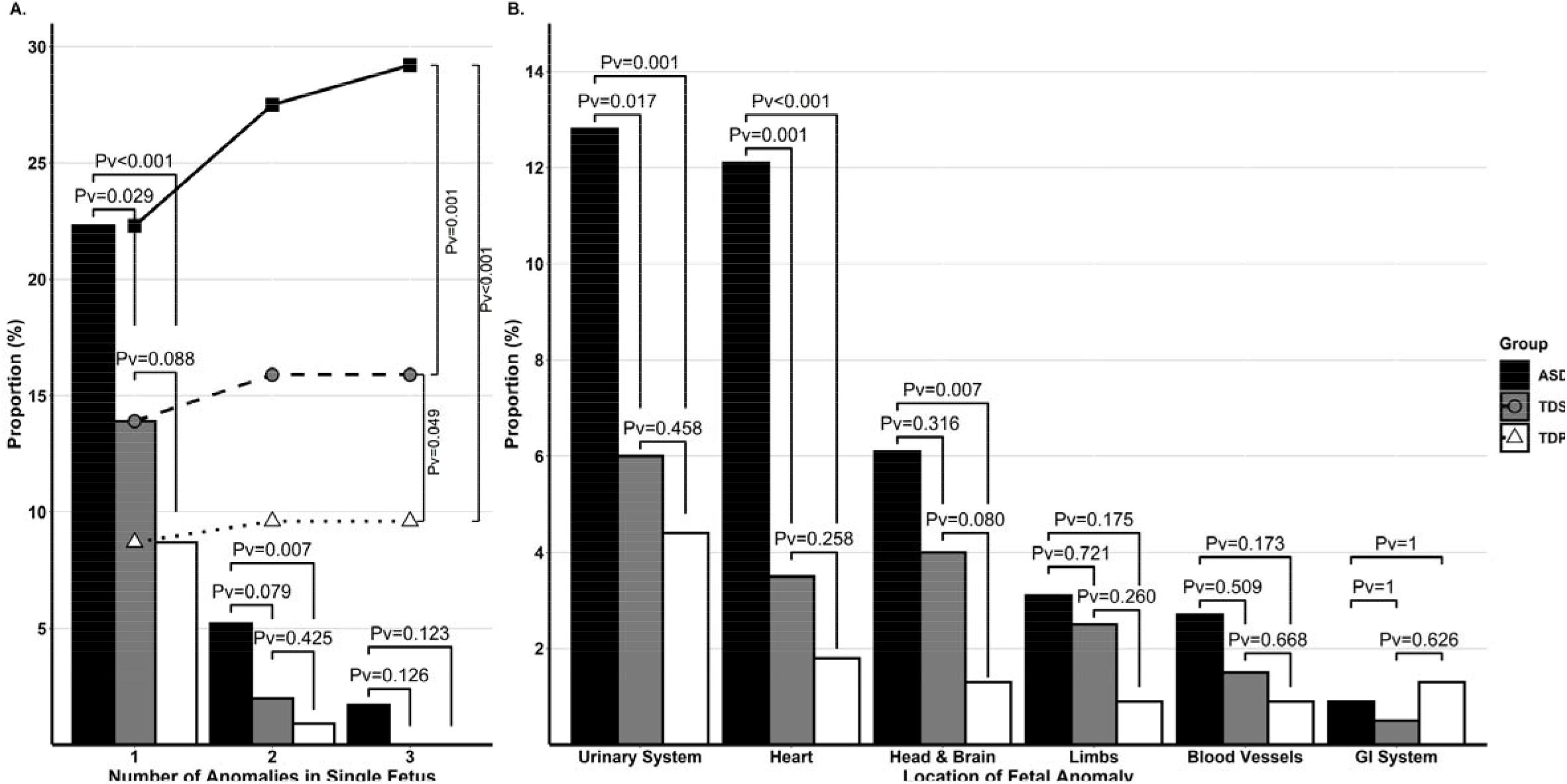
Proportions of anomalies in different fetal organ systems. (A.) Proportion of UFAs in each group. The bars represent the proportion of fetuses with 1,2, and 3 co-occurring UFAs. The lines with symbols represent the cumulative proportion of anomalies in each group: ASD (solid black line & black squares), TDS (dashed black line & gray circles), and TDP (dotted black line & white triangles). (B.) Proportion of UFAs in the different organs. Black bars –ASD; gray bars – TDS; white bars – TDP.

**Table 2.**
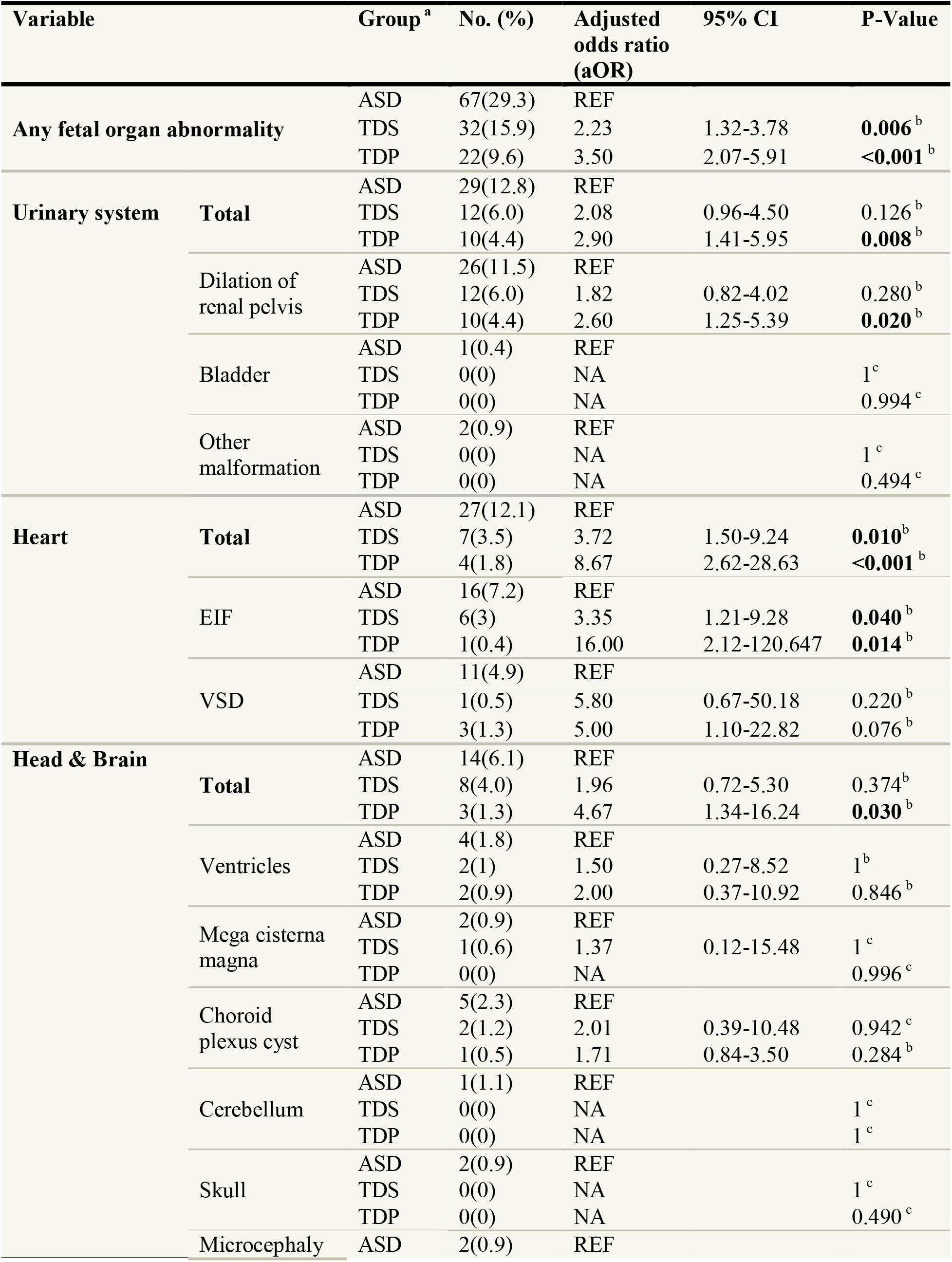
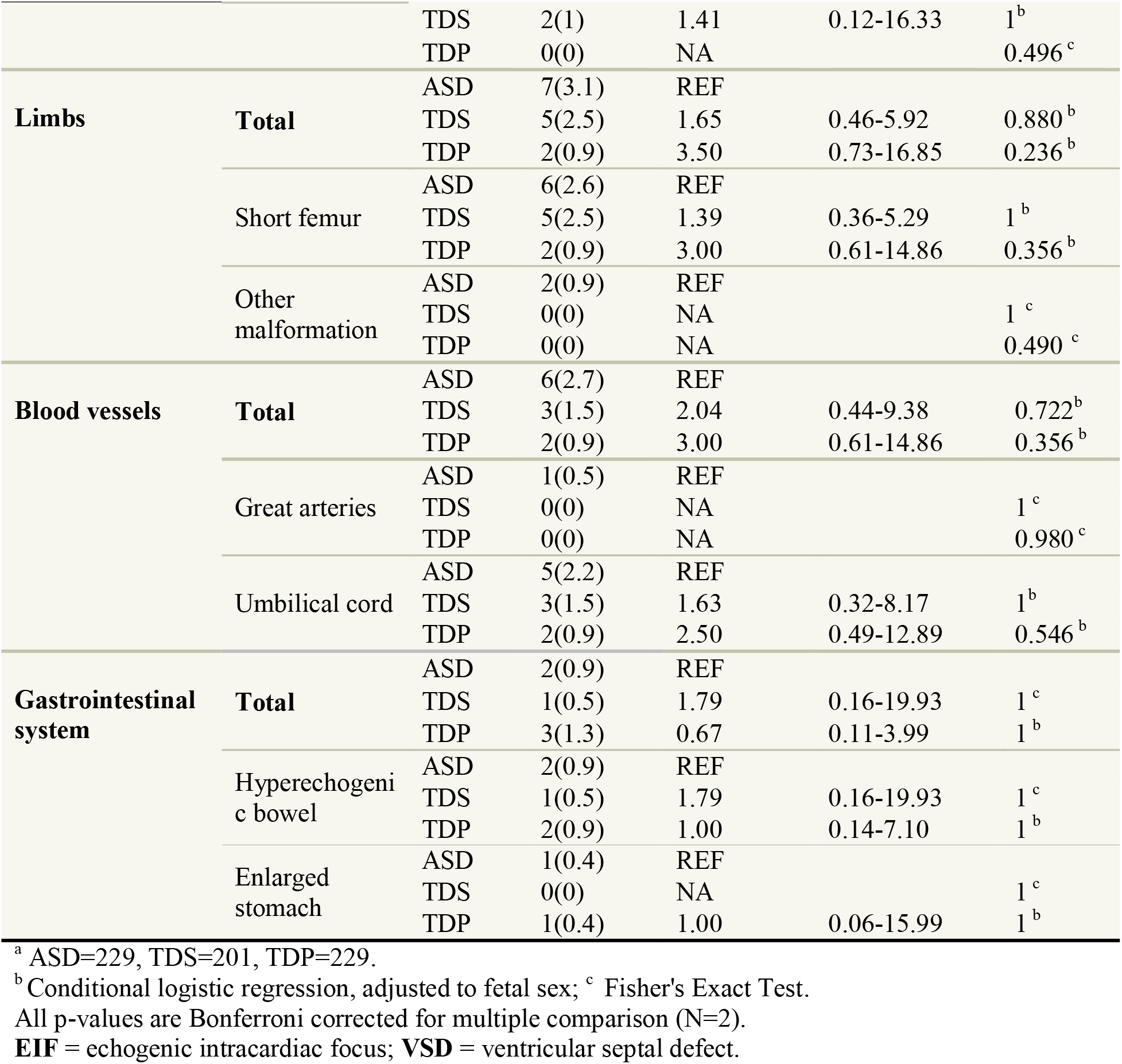
Anomalies in Different Fetal Organ Systems.

Common biometric measures evaluated during the anatomy survey are presented in **Table 3**. While ASD cases had significantly smaller (HC) and narrower (BPD) heads compared to TDP controls (aOR_HC_=0.76, 95%CI=0.62-0.93; aOR_BPD_=0.81, 95%CI=0.70-0.94), their ocular distance, relative to their BPD, was significantly larger than that in TDP controls (aOR_Ocular Distance_=1.29, 95%CI=1.06-1.57). ASD fetuses also had a lower biophysical profile than TDP fetuses (aOR=0.58, 95%CI=0.38-0.89), suggesting abnormal fetal neurodevelopment in the ASD cases. Finally, there were no significant differences in the sizes of the cisterna magna, cerebellum, or lateral ventricles between ASD fetuses and the two control groups.

**Table 3.** Risk of ASD Associated with Fetal Measures.

| Variable | Group | Mean ± SD | Odds Ratio (OR) | 95% CI | P-Value | Adjusted odds ratio (aOR) | 95% CI | P-Value |
| --- | --- | --- | --- | --- | --- | --- | --- | --- |
| zHC | ASD=227 | -0.11±1.0 | REF | | | | | |
| | TDS =199 | -0.20±1.1 | 1.08 | 0.90-1.30 | 0.782 | 0.98 | 0.71-1.19 | 1<sup>a</sup> |
| | TDP=227 | 0.12±0.8 | 0.77 | 0.63-0.94 | **0.020** | 0.76 | 0.62-0.93 | **0.018**<sup>a</sup> |
| zBPD | ASD=228 | 0.08±1.4 | REF | | | | | |
| | TDS =200 | -0.07±1.4 | 1.08 | 0.94-1.24 | 0.536 | 1.01 | 0.88-1.16 | 1<sup>a</sup> |
| | TDP=228 | 0.42±1.2 | 0.82 | 0.71-0.95 | **0.014** | 0.81 | 0.70-0.94 | **0.010**<sup>a</sup> |
| zAC | ASD=229 | -0.31±0.9 | REF | | | | | |
| | TDS =199 | -0.28±1.0 | 0.97 | 0.80-1.18 | 1 | 0.89 | 0.73-1.09 | 0.540<sup>a</sup> |
| | TDP=228 | -0.13±0.9 | 0.81 | 0.66-0.99 | 0.076 | 0.81 | 0.66-0.99 | 0.072<sup>a</sup> |
| zFL | ASD=229 | -0.20±0.9 | REF | | | | | |
| | TDS =200 | -0.13±0.8 | 0.91 | 0.73-1.14 | 0.838 | 0.89 | 0.70-1.12 | 0.606<sup>a</sup> |
| | TDP=227 | -0.15±0.8 | 0.93 | 0.74-1.16 | 1 | 0.93 | 0.74-1.16 | 1<sup>a</sup> |
| Ocular Distance, % | ASD=35 | 66.31±3.1 | REF | | | REF | | |
| | TDS =23 | 65.65±2.6 | 1.07 | 0.90-1.28 | 0.858 | 1.11 | 0.89-1.37 | 0.712<sup>b</sup> |
| | TDP=28 | 63.61±3.4 | 1.31 | 1.09-1.57 | **0.008** | 1.29 | 1.06-1.57 | **0.020**<sup>b</sup> |
| Cerebellum, % | ASD=91 | 45.53±2.6 | REF | | | REF | | |
| | TDS =69 | 45.20±2.3 | 1.06 | 0.93-1.20 | 0.818 | 1.04 | 0.91-1.19 | 1<sup>b</sup> |
| | TDP=81 | 45.01±2.6 | 1.08 | 0.96-1.22 | 0.380 | 1.08 | 0.96-1.22 | 0.282<sup>b</sup> |
| Cisterna Magna, mm | ASD=88 | 4.91±1.3 | REF | | | REF | | |
| | TDS =60 | 4.64±1.5 | 1.15 | 0.90-1.50 | 0.518 | 1.10 | 0.86-1.41 | 0.926<sup>b</sup> |
| | TDP=81 | 5.00±1.4 | 0.95 | 0.76-1.19 | 1 | 0.95 | 0.76-1.19 | 1<sup>b</sup> |
| Lateral Ventricles, mm | ASD=93 | 5.74±1.3 | REF | | | REF | | |
| | TDS =68 | 5.33±1.3 | 1.27 | 0.99-1.63 | 0.116 | 1.26 | 0.98-1.62 | 0.142<sup>b</sup> |
| | TDP=78 | 5.63±1.4 | 1.06 | 0.85-1.33 | 1 | 1.07 | 0.85-1.34 | 1<sup>b</sup> |
| Amniotic Fluid Index, cm | ASD=19 | 16.63±4.5 | REF | | | | | |
| | TDS =26 | 17.73±3.5 | 0.93 | 0.79-1.09 | 0.718 | 0.86 | 0.71-1.03 | 0.208<sup>b</sup> |
| | TDP=19 | 18.37±4.0 | 0.90 | 0.76-1.06 | 0.444 | 0.92 | 0.77-1.10 | 0.716<sup>b</sup> |
| Biophysical Profile, Score (1-8) | ASD=53 | 7.09±1.0 | REF | | | | | |
| | TDS =31 | 6.90±1.0 | 1.21 | 0.78-1.89 | 0.678 | 1.11 | 0.68-1.83 | 1<sup>b</sup> |
| | TDP=67 | 7.46±0.9 | 0.67 | 0.46-0.98 | 0.074 | 0.58 | 0.38-0.89 | **0.026**<sup>b</sup> |

<sup>a</sup> Conditional logistic regression, adjusted to fetal sex ; <sup>b</sup> logistic regression, adjusted to fetal sex and gestational age.
All p-values are Bonferroni corrected for multiple comparison (N=2).
**HC** = head circumference; **BPD** = biparietal diameter; **AC** = abdominal circumference; **FL** = femur length.

### Sex Differences

Sex differences in UFA rates are depicted in **Supplementary Table S2**. UFAs were significantly more common among ASD females than males (43.1% vs. 25.3%, *p*=0.013; for females and males respectively). The most significant sex differences were seen in UFAs in heart (23.5% vs. 8.7%, *p*=0.007), head&brain (11.8% vs. 4.5%, *p*=0.056), and gastrointestinal system (3.9% vs. 0%, *p*=0.051). In addition, ASD females had a higher prevalence of multiple co-occurring UFAs compared to ASD males (15.7% vs. 4.5%, *p*=0.011). TDS and TDP controls had no significant differences between females and males.

### Association with ASD Severity

Finally, we examined the association between UFAs and biometric measures with the severity of ASD symptoms (**Table 4**). The most significant associations were those between fetal cardiac anomalies and younger age at diagnosis (*p*=0.031), and between UFAs of the head&brain and DSM5-A criteria (*p*=0.017). Specifically, children for whom there were observable cardiac anomalies during gestation were diagnosed with ASD 6 months earlier than other children with ASD (33.6±11.7 vs. 39.4±16.5 months, *p*=0.031). Furthermore, children for whom there were observable head&brain UFAs were diagnosed as requiring more support than other children with ASD according to DSM5-A criteria.

**Table 4.**
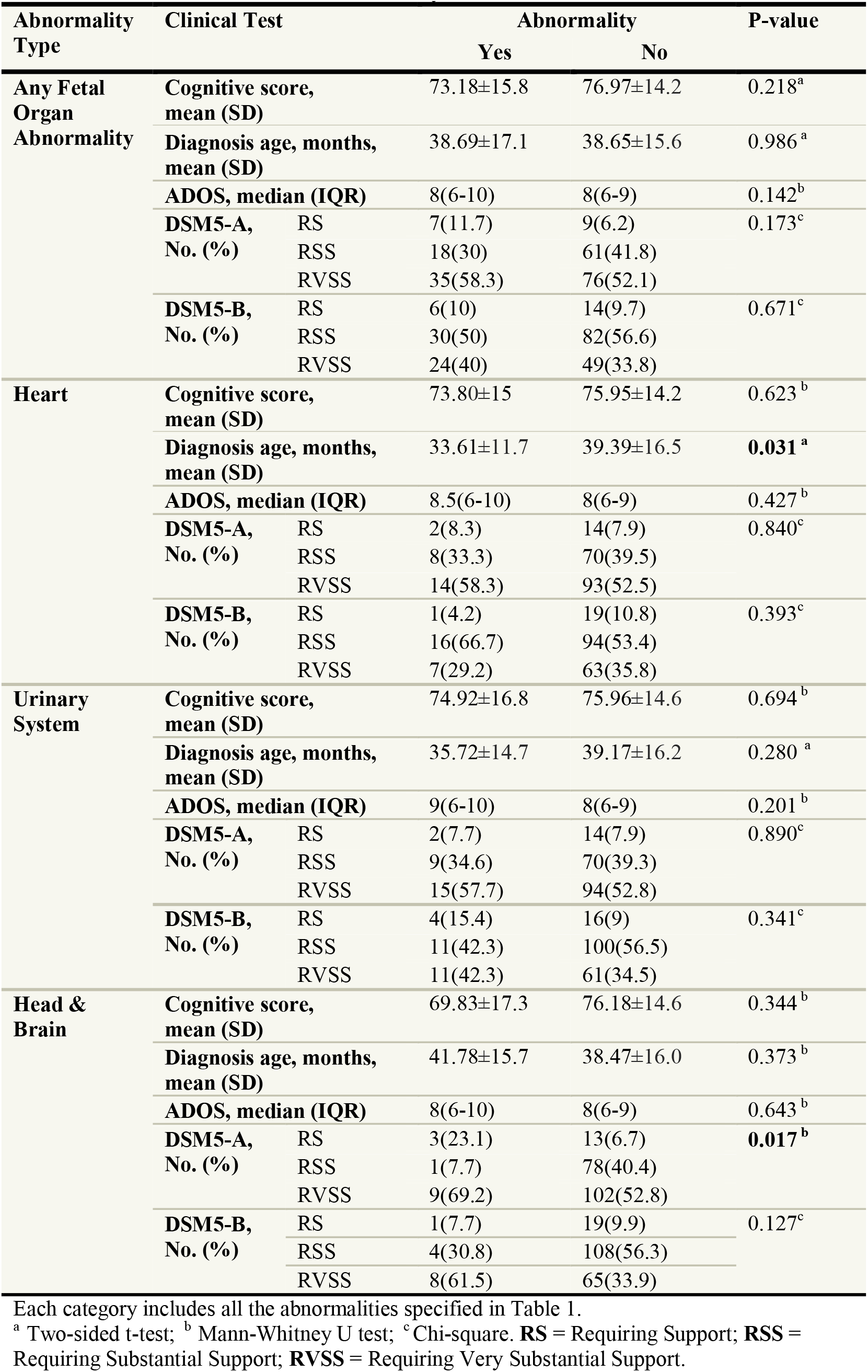
Association between Clinical Severity and Fetal Abnormalities.

### Sensitivity analysis

The study sample included one randomly selected ASD case from each of the eight multiplex families in the study. We repeated all the reported analyses in this study using a sample that included the other ASD case in each family. The results of these analyses are reported in **Supplementary Tables S3-S5** and show the same differences in UFA rates between the study groups.

## Discussion

This study is the first to comprehensively examine prenatal organ development in ASD children via an examination of the fetal anatomy survey. We show that fetuses developing into children later diagnosed with ASD had significantly higher rates of UFAs compared to both their typically developing siblings and to matched typically developing children from the general population. These finding highlight the association of certain UFAs with ASD susceptibility of the developing fetus. These UFAs, which can be detected in standard prenatal anatomy ultrasound surveys conducted during mid-gestation, could form the basis of new prenatal screening approaches for ASD. The results of such prenatal screening will reveal fetuses at risk to develop ASD and may facilitate their earlier diagnosis, a factor that has already been shown to optimize the long-term outcomes of ASD treatment.^53–55^

Most of the identified UFAs were observed in the urinary system, heart, and head&brain, suggesting a shared etiology for the abnormal development of these organs in ASD. Dilation of the renal pelvis (pyelectasis or hydronephrosis) was the most prevalent UFA among ASD cases in our study. This finding is consistent with the higher rates of pyelectasis in ASD fetuses compared with the prevalence of this anomaly in the general population.^27^ Pyelectasis is considered a “soft marker” associated with an underlying fetal genetic risk.^37,39^ Furthermore, a number of genetic syndromes associated with ASD are characterized by various renal anomalies. For example, children with the 16q24.2 deletion or the17q12 microdeletion usually manifest both ASD and various congenital abnormalities of the kidney and urinary tract, including dilation of the renal pelvis,^31,32,56–58^; some of these congenital abnormalities could indeed be identified in prenatal ultrasound scans.^28–30,32^ Another example is Phelan-McDermid syndrome, caused by 22q13 deletion or by disruptive mutations in *SHANK3*, one of the most common monogenic causes of ASD, with renal abnormalities being found in 25–38% of children with this syndrome.^11,59^

As mentioned above, higher odds of an ASD diagnosis were significantly associated with cardiac UFAs, including echogenic intracardiac focus, which is considered a “soft marker” associated with various genetic anomalies,^37^ and ventricular septal defect, which is a structural malformation that may progress to congenital heart disease (CHD) after birth.^4,34^ Indeed, there is emerging evidence supporting a possible association between CHD and ASD, with several population-based studies reporting a higher risk of ASD in children with CHD.^4–6,9,13,16,60–62^ Furthermore, recent findings from exome sequencing studies demonstrate a striking overlap between genes associated with ASD and CHD.^4,63^ A genetic link between ASD and CHD can also be seen in several genetic syndromes associated with ASD; for example, approximately 3– 25% of children with Phelan-McDermid syndrome also manifest various cardiac abnormalities,^11,59^ and comorbidity of ASD and cardiac defects is also seen in children with the 22q11 deletion.^64,65^

Relatively high UFA rates were also seen for the head&brain. These UFAs consisted mainly of anomalies in the cerebrospinal fluid (CSF) circulation, including choroid plexus cysts, enlarged lateral ventricles, and mega cisterna magna, suggesting abnormal development of CSF circulation in ASD compared to TDP. Indeed, increased pre- and postnatal ventricle volumes have been proposed as early structural markers of altered development of the cerebral cortex and increased risk for neuropsychiatric disorders, including ASD.^23,26,66^ In addition, ventriculomegaly, enlarged cisterna magna, hydrocephalus, and increased extra-axial CSF were associated with ASD in multiple MRI and population-based studies.^7,23,26,66–72^ Finally, children with 22q13 deletion syndrome associated with ASD are characterized by abnormalities in the CSF circulation, including ventricle dilation, enlarged cisterna magna, and arachnoid cysts.^11,59,73^

Abnormalities in the CSF circulation in the extra-axial space may lead to an accumulation of CSF above the frontal lobes,^69–72^ resulting in an abnormal and elongated (dolichocephalic) head shape, as revealed in this analysis and our previous smaller study.^21^ In addition, ASD fetuses had relatively wider set eyes vs. the other fetuses in the cohort. This finding is in line with evidence from postnatal head image analysis demonstrating wide-set eyes in a subgroup of children with ASD.^74^ Both dolichocephaly and wide-set eyes have been linked to several genetic anomalies associated with ASD, including copy-number variants in the 16p11.2^75^ and 22q13^11,76,77^ chromosomal loci and mutations in the *CHD8* gene.^78^

Our findings also suggest a positive association between fetal structural anomalies and ASD severity. Indeed, congenital anomalies have been shown to be more prevalent among individuals with autism and intellectual disability,^12^ and ASD children with CHD or wider-set eyes have worse cognitive, language, and attention disabilities than other children with ASD.^13,61,74^ In addition, ASD children with CHD usually also suffer from developmental delay and tend to be diagnosed earlier than most other children with ASD.^61^ Furthermore, the amount of extra-axial CSF volume detected as early as 6 months is predictive of more severe ASD symptoms.^71,72^ Finally, children with genetic syndromes that include both ASD and congenital malformations usually manifest additional cognitive and clinical impairments that lead to a more severe ASD outcome.^31,64,65^

We show that ASD females have more UFAs and multiple co-occurring UFAs compared to ASD males. These findings are in line with the higher prevalence of comorbidities, including congenital anomalies, among ASD females,^6,8,79^ and with our previous report about sex differences in prenatal head growth in children with ASD^21^. These findings are also in line with the reported higher prevalence of genetic abnormalities in ASD females compared to ASD males,^79–83^ and with the known more severe manifestation of ASD in females.^8,79,84^ Altogether, these evidence are consistent with theories about diverged etiologies of ASD in males and females.^79,85,86^

This study is the first to systematically examine organogenesis in fetuses later developing into children with ASD by exploiting retrospectively the fetal ultrasound anatomy survey. The use of two distinct control groups, TDS and TDP, enabled us to adjust our findings to multiple familial and prenatal confounders that are known to have a considerable effect on both ASD risk and fetal growth (e.g., sex^2,21^ and shared genetics among siblings^1,87,88^), making our findings more compelling.

### Study Limitations

The study has some limitations. There was a slight underrepresentation of Jews in the study sample, probably because Jewish parents tend to use private insurance to conduct a more comprehensive anatomy survey than that offered by the HMO. In addition, despite the large size of the study cohort, consisting of over 650 children, it still lacked sufficient statistical power to enable us to draw conclusions about rare UFAs (e.g., UFAs in the cerebellum, cisterna magna, great arteries, and gastrointestinal system) or about variables with a significant fraction of missing data such as biometric measures and clinical severity.

## Conclusions

The association of UFAs with ASD, especially in the urinary system, heart, head, and brain, sheds important light on the abnormal multiorgan embryonic development of this complex disorder and suggests several fetal ultrasonography biomarkers for ASD.

## Data Availability

Data referred to in the manuscript is confidential

## Acknowledgements

We thank Mrs. Inez Mureinik for critical reviewing and editing of the manuscript.

This study was conducted as part of the requirements to obtain a degree in medicine from the Joyce & Irving Goldman Medical School, Faculty of Health Sciences, Ben-Gurion University of the Negev.

The article has been previously posted in MedRxiv preprint server.

## Funding

This study was supported by a grant from the Israeli Science Foundation (1092/21).

## Competing Interests

The authors report no biomedical financial interests or potential conflicts of interest.

## Notes

### Competing Interest Statement

The authors have declared no competing interest.

### Author Declarations

The study was approved by the Ethics Committee of the Soroka University Medical Center, Beer-Sheva, Israel per the Helsinki declaration (SOR #295-18).

### Summary of Updates

Addition of analysis on sex differencecs

