## Supplementary Tables for "Association Between Ultrasonography Fetal Anomalies and Autism Spectrum Disorder"

### Supplementary Appendix

**Supplementary Table S1.** Clinical and Sociodemographic Characteristics of ASD Children

**Supplementary Table S2.** Sex Differences in Prevalence of UFAs

**Supplementary Table S3.** Anomalies in Different Fetal Organ Systems – Sensitivity Analysis

**Supplementary Table S4.** Risk of ASD Associated with Fetal Measures – Sensitivity Analysis

**Supplementary Table S5.** Association between Clinical Severity and Fetal Abnormalities – Sensitivity Analysis

| Supplementary Table S1. Clinical and Sociodemographic Characteristics of ASD Children | | | |
|---|---|---|---|
| **Variable** | **Included (n = 229)** | **Excluded (n = 475)** | **P-Value** |
| Jewish, no. (%) [a] | 159(70.4) | 387 (81.5) | **0.001** |
| Male, no. (%) [a] | 179(78.5) | 370(77.8) | 0.820 |
| Moderate salary or lower, No. (%) [a] | 38(60.3) | 96(64.4) | 0.570 |
| Special education setting, No. (%) [a] | 59(30.4) | 124(34.8) | 0.147 |
| Mother's age, mean±SD, y [b] | 29.6±5.5 | 31.7±6.2 | **<0.001** |
| Father's age, mean±SD, y [b] | 32.8±7.8 | 34.8±7.4 | **0.003** |
| Diagnosis age, mean±SD, y [b] | 3.2±1.3 | 3.2±1.5 | 0.666 |
| Cognitive score (IQ), mean±SD [b] | 75.8±14.8 | 76.3±17.1 | 0.789 |
| ADOS Comparison Score, median (IQR) [c] | 8(6-9) | 7(6-9) | **0.009** |

[a] Chi-square; [b] Two-sided t-test; [c] Mann-Whitney U test

| Supplementary Table S2. Sex Differences in Prevalence of UFAs | | | | |
|---|---|---|---|---|
| **Variable** | **Group** | **Male no. (%)** | **Female no. (%)** | **P-value[a]** |
| **Any Fetal Organ Abnormality** | ASD | 45(25.3) | 22(43.1) | **0.013** |
| | TDS | 15(13.2) | 17(19.5) | 0.220 |
| | TDP | 19(10.7) | 3(5.9) | 0.423 |
| **Urinary System** | ASD | 22(12.5) | 7(13.7) | 0.817 |
| | TDS | 8(7) | 4(4.6) | 0.473 |
| | TDP | 10(5.6) | 0(0) | 0.123 |
| **Heart** | ASD | 15(8.7) | 12(23.5) | **0.007** |
| | TDS | 2(1.8) | 5(5.7) | 0.132 |
| | TDP | 2(1.1) | 2(4) | 0.213 |
| **Head & Brain** | ASD | 8(4.5) | 6(11.8) | 0.056 |
| | TDS | 4(3.5) | 4(4.6) | 0.696 |
| | TDP | 3(1.7) | 0(0) | 0.351 |
| **Limbs** | ASD | 6(3.4) | 1(2) | 1 |
| | TDS | 1(0.9) | 4(4.6) | 0.168 |
| | TDP | 2(1.1) | 0(0) | 1 |
| **Blood Vessels** | ASD | 3(1.7) | 3(5.9) | 0.132 |
| | TDS | 2(1.8) | 1(1.1) | 1 |
| | TDP | 1(0.6) | 1(2) | 0.397 |
| **Gastrointestinal System** | ASD | 0(0) | 2(3.9) | 0.051 |
| | TDS | 0(0) | 1(1.1) | 0.433 |
| | TDP | 0(0) | 3(1.3) | 1 |
| **UFA in >1 organ** | ASD | 8(4.5) | 8(15.7) | **0.011** |
| | TDS | 1(0.9) | 3(3.4) | 0.217 |
| | TDP | 2(1.1) | 0(0) | 0.603 |

[a] Chi-Square

UFA = Ultrasonography Fetal Anomaly

**Supplementary Table S3. Anomalies in Different Fetal Organ Systems – Sensitivity Analysis**

| Variable | | Group [a] | No. (%) | Adjusted odds ratio (aOR) | 95% CI | P-Value |
|---|---|---|---|---|---|---|
| **Any Fetal Organ Abnormality** | | ASD | 65(28.4) | REF | | |
| | | TDS | 32(15.9) | 2.11 | 1.25-3.55 | **0.010** [b] |
| | | TDP | 24(10.5) | 3.16 | 1.89-5.29 | **<0.001** [b] |
| **Urinary System** | **Total** | ASD | 28(12.2) | REF | | |
| | | TDS | 12(6.0) | 2.03 | 0.93-4.41 | 0.150 [b] |
| | | TDP | 10(4.4) | 2.80 | 1.36-5.76 | **0.010** [b] |
| | Dilation of Renal Pelvis | ASD | 25(10.9) | REF | | |
| | | TDS | 12(6.0) | 1.77 | 0.79-3.93 | 0.326 [b] |
| | | TDP | 10(4.4) | 2.50 | 1.20-5.21 | **0.028** [b] |
| | Bladder | ASD | 1(0.4) | REF | | |
| | | TDS | 0(0) | NA | | 1 [c] |
| | | TDP | 0(0) | NA | | 0.994 [c] |
| | Other Malformation | ASD | 2(0.9) | REF | | |
| | | TDS | 0(0) | NA | | 1 [c] |
| | | TDP | 0(0) | NA | | 0.494 [c] |
| **Heart** | **Total** | ASD | 26(11.4) | REF | | |
| | | TDS | 7(3.5) | 3.55 | 1.42-8.88 | **0.014** [b] |
| | | TDP | 5(2.2) | 6.25 | 2.18-17.96 | **0.002** [b] |
| | EIF | ASD | 16(7.0) | REF | | |
| | | TDS | 6(3) | 3.35 | 1.21-9.28 | **0.040** [b] |
| | | TDP | 2(0.9) | 8.00 | 1.84-34.79 | **0.012** [b] |
| | VSD | ASD | 10(4.4) | REF | | |
| | | TDS | 1(0.5) | 4.76 | 0.53-42.97 | 0.330 [b] |
| | | TDP | 3(1.3) | 4.50 | 0.97-20.83 | 0.108 [b] |
| **Head & Brain** | **Total** | ASD | 13(5.7) | REF | | |
| | | TDS | 8(4.0) | 1.67 | 0.64-4.33 | 0.584 [b] |
| | | TDP | 3(1.3) | 4.33 | 1.24-15.21 | **0.044** [b] |
| | Ventricles | ASD | 4(1.7) | REF | | |
| | | TDS | 2(1) | 1.50 | 0.27-8.52 | 1 [b] |
| | | TDP | 2(0.9) | 2.00 | 0.37-10.92 | 0.846 [b] |
| | Mega Cisterna Magna | ASD | 2(0.9) | REF | | |
| | | TDS | 1(0.5) | 2.79 | 0.24-32.70 | 0.828 [b] |
| | | TDP | 0(0) | NA | | 0.996 [c] |
| | Choroid Plexus Cyst | ASD | 4(1.7) | REF | | |
| | | TDS | 2(1.0) | 1.71 | 0.81-3.58 | 0.318 [b] |
| | | TDP | 1(0.4) | 1.59 | 0.77-3.30 | 0.430 [b] |
| | Cerebellum | ASD | 0(0) | REF | | |
| | | TDS | 0(0) | NA | | 1 [c] |
| | | TDP | 0(0) | NA | | 1 [c] |
| | Skull | ASD | 2(0.9) | REF | | |
| | | TDS | 0(0) | NA | | 1 [c] |
| | | TDP | 0(0) | NA | | 0.490 [c] |
| | Microcephaly | ASD | 2(0.9) | REF | | |
| | | TDS | 2(1) | 1.41 | 0.12-16.33 | 1 [b] |
| | | TDP | 0(0) | NA | | 0.496 [c] |
| **Limbs** | **Total** | ASD | 7(3.1) | REF | | |
| | | TDS | 5(2.5) | 1.65 | 0.46-5.92 | 0.880 [b] |
| | | TDP | 2(0.9) | 3.50 | 0.73-16.85 | 0.236 [b] |
| | Short Femur | ASD | 6(2.6) | REF | | |
| | | TDS | 5(2.5) | 1.39 | 0.36-5.29 | 1 [b] |
| | | TDP | 2(0.9) | 3.00 | 0.61-14.86 | 0.356 [b] |
| | Other Malformation | ASD | 2(0.9) | REF | | |

| | | | | | | |
|---|---|---|---|---|---|---|
| | | TDS | 0(0) | NA | | 1 [c] |
| | | TDP | 0(0) | NA | | 0.490 [c] |
| **Blood Vessels** | | ASD | 6(2.7) | REF | | |
| | **Total** | TDS | 3(1.5) | 2.04 | 0.44-9.38 | 0.722 [b] |
| | | TDP | 2(0.9) | 3.00 | 0.61-14.86 | 0.356[b] |
| | | ASD | 1(0.5) | REF | | |
| | Great Arteries | TDS | 0(0) | NA | | 1 [c] |
| | | TDP | 0(0) | NA | | 0.980 [c] |
| | | ASD | 5(2.2) | REF | | |
| | Umbilical Cord | TDS | 3(1.5) | 1.63 | 0.32-8.17 | 1 [b] |
| | | TDP | 2(0.9) | 2.50 | 0.49-12.89 | 0.546[b] |
| | | ASD | 2(0.9) | REF | | |
| | **Total** | TDS | 1(0.5) | 1.79 | 0.16-19.93 | 1 [c] |
| | | TDP | 3(1.3) | 0.67 | 0.11-3.99 | 1 [b] |
| **Gastrointestinal System** | Hyperechogenic bowel | ASD | 2(0.9) | REF | | |
| | | TDS | 1(0.5) | 1.79 | 0.16-19.93 | 1 [c] |
| | | TDP | 2(0.9) | 1.00 | 0.14-7.10 | 1[b] |
| | | ASD | 1(0.4) | REF | | |
| | Enlarged stomach | TDS | 0(0) | NA | | 1 [c] |
| | | TDP | 1(0.4) | 1.00 | 0.06-15.99 | 1[b] |

[a] ASD=229, TDS=201, TDP=229.
[b] Conditional logistic regression, adjusted to fetal sex; [c] Fisher's Exact Test.
All p-values are Bonferroni corrected for multiple comparison (N=2).
**EIF** = echogenic intracardiac focus; **VSD** = ventricular septal defect.

**Supplementary Table S4. Risk of ASD Associated with Fetal Measures – Sensitivity Analysis**

| Variable | Group | Mean ± SD | Odds Ratio (OR) | 95% CI | P-Value | Adjusted odds ratio (aOR) | 95% CI | P-Value |
|---|---|---|---|---|---|---|---|---|
| zHC | ASD=227 | -0.10±1.0 | REF | | | REF | | |
| | TDS =199 | -0.20±1.1 | 1.10 | 0.91-1.32 | 0.652 | 0.96 | 0.77-1.21 | 1 [a] |
| | TDP=227 | 0.14±1.1 | 0.79 | 0.65-0.96 | **0.030** | 0.78 | 0.64-0.96 | **0.040** [a] |
| zBPD | ASD=228 | 0.10±1.4 | REF | | | REF | | |
| | TDS =200 | -0.07±1.4 | 1.09 | 0.95-1.25 | 0.436 | 1.05 | 0.88-1.25 | 1[a] |
| | TDP=228 | 0.46±1.4 | 0.83 | 0.73-0.96 | **0.018** | 0.83 | 0.72-0.96 | **0.026** [a] |
| zAC | ASD=229 | -0.29±0.9 | REF | | | REF | | |
| | TDS =199 | -0.28±1.0 | 0.99 | 0.82-1.21 | 1 | 0.92 | 0.73-1.16 | 0.980 [a] |
| | TDP=228 | -0.12±1.0 | 0.84 | 0.70-1.02 | 0.152 | 0.84 | 0.68-1.02 | 0.156 [a] |
| zFL | ASD=229 | -0.20±0.9 | REF | | | REF | | |
| | TDS =200 | -0.13±0.8 | 0.92 | 0.73-1.15 | 0.884 | 0.86 | 0.64-1.14 | 0.584 [a] |
| | TDP=227 | -0.14±0.9 | 0.92 | 0.75-1.14 | 0.898 | 0.93 | 0.75-1.15 | 0.968 [a] |
| Ocular Distance, % | ASD=35 | 66.44±3.5 | REF | | | REF | | |
| | TDS =23 | 65.65±2.6 | 1.08 | 0.91-1.29 | 0.720 | 1.11 | 0.90-1.37 | 0.668[b] |
| | TDP=28 | 63.77±3.1 | 1.28 | 1.08-1.52 | **0.010** | 1.25 | 1.04-1.50 | **0.034**[c] |
| Cerebellum, % | ASD=91 | 45.53±2.6 | REF | | | REF | | |
| | TDS =69 | 45.20±2.3 | 1.05 | 0.93-1.20 | 0.820 | 1.04 | 0.91-1.19 | 1 [b] |
| | TDP=80 | 45.06±2.7 | 1.07 | 0.95-1.20 | 0.512 | 1.08 | 0.96-1.21 | 0.432 [b] |
| Cisterna Magna, mm | ASD=89 | 4.88±1.3 | REF | | | REF | | |
| | TDS =60 | 4.64±1.5 | 1.13 | 0.89-1.44 | 0.622 | 1.08 | 0.84-1.39 | 1 [b] |
| | TDP=80 | 4.97±1.3 | 0.95 | 0.75-1.19 | 1 | 0.95 | 0.75-1.19 | 1 [b] |
| Lateral Ventricles, mm | ASD=92 | 5.72±1.3 | REF | | | REF | | |
| | TDS =68 | 5.33±1.3 | 1.26 | 0.98-1.61 | 0.136 | 1.24 | 0.97-1.60 | 0.180 [b] |
| | TDP=80 | 5.55±1.4 | 1.10 | 0.88-1.37 | 0.808 | 1.11 | 0.88-1.34 | 0.748 [b] |
| Amniotic Fluid Index, cm | ASD=20 | 16.60±4.4 | REF | | | | | |
| | TDS =26 | 17.73±3.5 | 0.92 | 0.78-1.09 | 0.670 | 0.85 | 0.70-1.03 | 0.180 [b] |
| | TDP=18 | 18.56±4.1 | 0.90 | 0.75-1.05 | 0.344 | 0.89 | 0.77-1.08 | 0.516 [b] |
| Biophysical Profile, Score (1-8) | ASD=53 | 7.13±1.0 | REF | | | | | |
| | TDS =31 | 6.90±1.0 | 1.26 | 0.81-1.97 | 0.624 | 1.17 | 0.70-193 | 1 [b] |
| | TDP=68 | 7.47±0.9 | 0.69 | 0.47-1.01 | 0.106 | 0.62 | 0.41-0.94 | **0.050** [b] |

[a] Conditional logistic regression, adjusted to fetal sex; [b] logistic regression, adjusted to fetal sex and gestational age.
All p-values are Bonferroni corrected for multiple comparison (N=2).
**HC** = head circumference; **BPD** = biparietal diameter; **AC** = abdominal circumference; **FL** = femur length.

**Supplementary Table S5. Association between Clinical Severity and Fetal Abnormalities – Sensitivity Analysis**

| Abnormalities Type | Clinical Test | | Abnormality | No Abnormality | P-Value |
|---|---|---|---|---|---|
| **Any Fetal Organ Abnormality** | **Cognitive Score, mean (SD)** | | 73.18±15.8 | 76.62±14.1 | 0.243[a] |
| | **Diagnosis Age, Months, mean (SD)** | | 38.53±17.0 | 37.64±15.4 | 0.986[a] |
| | **ADOS, median (IQR)** | | 8(6-10) | 8(6-9) | 0.259[b] |
| | **DSM5-A, No. (%)** | RS | 7(12.1) | 9(6.2) | 0.116[c] |
| | | RSS | 16(27.6) | 60(41.1) | |
| | | RVSS | 35(60.3) | 77(52.7) | |
| | **DSM5-B, No. (%)** | RS | 5(8.6) | 14(9.6) | 0.609[c] |
| | | RSS | 28(48.3) | 80(54.8) | |
| | | RVSS | 25(43.1) | 52(35.6) | |
| **Heart** | **Cognitive Score, mean (SD)** | | 73.80±15 | 75.58±14.2 | 0.700[b] |
| | **Diagnosis Age, Months, mean (SD)** | | 33.31±11.8 | 38.51±16.3 | 0.056[a] |
| | **ADOS, median (IQR)** | | 8 (6-10) | 8(6-9) | 0.489[b] |
| | **DSM5-A, No. (%)** | RS | 2(8.7) | 14(8.0) | 0.953[c] |
| | | RSS | 8(34.8) | 67(38.1) | |
| | | RVSS | 13(56.5) | 95(54) | |
| | **DSM5-B, No. (%)** | RS | 1(4.3) | 18(10.2) | 0.416[c] |
| | | RSS | 15(65.2) | 91(51.7) | |
| | | RVSS | 7(30.4) | 67(38.1) | |
| **Urinary System** | **Cognitive Score, mean (SD)** | | 74.50±17.5 | 75.66±14.5 | 0.660[b] |
| | **Diagnosis Age, Months, mean (SD)** | | 35.54±14.9 | 38.29±16.0 | 0.392[a] |
| | **ADOS, median (IQR)** | | 9(6-10) | 8(6-9) | 0.243[b] |
| | **DSM5-A, No. (%)** | RS | 2(8.0) | 14(7.9) | 0.818[c] |
| | | RSS | 8(32.0) | 68(38.4) | |
| | | RVSS | 15(60.0) | 95(53.7) | |
| | **DSM5-B, No. (%)** | RS | 3(12.0) | 16(9.0) | 0.623[c] |
| | | RSS | 11(44.0) | 96(54.2) | |
| | | RVSS | 11(44.0) | 65(36.7) | |
| **Head & Brain** | **Cognitive Score, mean (SD)** | | 69.83±17.3 | 75.86±14.6 | 0.369[b] |
| | **Diagnosis Age, Months, mean (SD)** | | 41.78±15.7 | 37.65±15.8 | 0.288[b] |
| | **ADOS, median (IQR)** | | 8(6-10) | 8(6-9) | 0.561[b] |
| | **DSM5-A, No. (%)** | RS | 3(23.1) | 13(6.8) | **0.019**[c] |
| | | RSS | 1(7.7) | 75(39.3) | |
| | | RVSS | 9(69.2) | 103(53.9) | |
| | **DSM5-B, No. (%)** | RS | 1(7.7) | 18(9.4) | 0.182[c] |
| | | RSS | 4(30.8) | 104(54.5) | |
| | | RVSS | 8(61.5) | 69(36.1) | |

Each category includes all the abnormalities specified in Table 1.
[a] Two-sided t-test; [b] Mann-Whitney U test; [c] Chi-square.
**RS** = Requiring Support; **RSS** = Requiring Substantial Support; **RVSS** = Requiring Very Substantial Support.
